# ‘Admissions to paediatric medical wards with a primary mental health diagnosis: a systematic review of the literature’

**DOI:** 10.1101/2023.11.10.23298368

**Authors:** Adriana Vázquez-Vázquez, Abigail Smith, Faith Gibson, Helen Roberts, Gabrielle Mathews, Joseph Ward, Russell Viner, Dasha Nicholls, Francesca Cornaglia, Damian Roland, Kirsty Phillips, Lee Hudson

## Abstract

**Objective:** To systematically review the literature describing children and young people (CYP) admissions to paediatric general wards because of primary mental health reasons, particularly in MH crisis.

**Design:** PubMed, Embase, PsychINFO, Web of Science and Google Scholar were searched. We addressed five search questions to inform: trends and/or the number of admissions, the risk factors for adverse care, the experiences of CYP, families/carers and health care professionals (HCPs) and the evidence of interventions aimed at improving the care during admissions.

Two reviewers independently assessed the relevance of abstracts identified, extracted data and undertook quality assessment. This review was registered with Prospero (CRD42022350655).

**Results:** Thirty-two studies met the inclusion criteria. Eighteen addressed trends and or numbers/proportions of admissions, 12 provided data about the views/experiences of HCPs, two provided data about CYPs’ experiences and four explored improving care. We were unable to identify studies examining risk factors for harm during admissions, but studies did report the length of stay in general paediatric/adult settings while waiting for specialised care, which could be considered a risk factor while caring for this group.

**Conclusions:** MH admissions to children’s wards are a long-standing issue and are increasing. CYP will continue to need to be admitted in crisis, with paediatric wards a common allocation whilst waiting for assessment. For services to be delivered effectively and for CYPs and their families/carers to feel supported and HCPs to feel confident, we need to facilitate more integrated physical and MH pathways of care.

**What is already known on this topic?:**

- There is evidence that both the number of paediatric admissions and the severity of MH crisis in CYP have increased. Children’s wards are not designed to treat CYP with MH problems and HCPs haven’t had enough training.
- There is no published systematic review on this topic.

**What this study adds?:**

- This is the first systematic review on CYP admissions to paediatric wards with a primary MH indication.
- Evidence suggested increased numbers of admissions to paediatric wards over time. HCPs reported concerns about skill sets to manage CYP with MH presentations and questioned the appropriateness of the acute ward for this care.
- There is limited evidence on CYP experiences during admissions and on studies to improve care. We found no evidence of specific risk factors for adverse care for CYP and families/carers during admissions and no studies on families/carers’ experiences.

## INTRODUCTION

Mental health (MH) disorders represent a significant burden on the health of children and young people (CYP) (1) with some CYP admitted to hospital because of a deterioration in their MH (2). In an emergency, such admissions tend to be to medical children’s wards (3) which may serve as an acute place for safety/assessment (4) or provide interventions such as treatment for overdose (5) or nutritional rehabilitation (6). Paediatric wards can also be a place of admission while waiting for a specialist MH admission, sometimes called “psychiatric boarding/psychiatric boarders (PBs)” (7,8). Although CYP with acute MH presentations have always formed part of the caseload of paediatric medical wards (3), clinicians are reporting that these admissions are becoming more common and more complex since the SARS-CoV-2 pandemic (7,9,10). MH admissions to paediatric wards present challenges for service users and providers alike. Paediatric wards may not be safely prepared for the numbers or specialist care needed (3).

A number of systematic reviews have also found limited efficacy for interventions to reduce admissions of CYP with a MH crisis (2,11), and there is evidence that CYP admitted with a MH diagnosis are more likely to require readmission (12). Such admissions are not just considerations for providing care on paediatric medical wards in the here and now but are likely to remain so for the foreseeable future. This calls for a focus on the quality and safety of care for such admissions for CYP, families/carers and the teams caring for them (13) to which an up-to-date synthesis of the published literature can contribute. Whilst a number of systematic reviews have focused on the care of CYP presenting to emergency departments (ED) with MH disorders (14–16), at the time of writing we were unable to find any systematic reviews on admissions to paediatric wards. Our broad systematic review of the literature on acute MH admissions to paediatric medical wards was carried out using PRISMA guidelines. We asked five questions: 1)To inform the size of the problem, what is the evidence for trends in the number of admissions and/or the number/proportions of CYP admitted to paediatric or adult wards because of a primary MH diagnosis?; 2) To inform factors that can impact care, what are the risk factors for poor care for CYP and families/carers during admissions to paediatric wards (or adult general wards) because of a primary MH diagnosis?; 3) To examine the context of care, what are the reported experiences of HCPs on paediatric wards (or adult general wards) during the admissions of CYP because of a primary MH diagnosis?; 4) To understand CYP and families/carers’ experiences as part of the context of care, what are the reported experiences of CYP and their families/carers during admissions to paediatric wards (or adult general wards) because of a primary MH diagnosis?; 5) To inform about support during MH admissions, is there evidence of interventions or quality improvement projects aimed at improving the care of CYP and families/carers during admissions to paediatric wards (or adult wards) because of a primary MH diagnosis?

## METHODS

### Protocol and registration

Our review protocol was registered with PROSPERO registry of systematic reviews (CRD42022350655).

### Eligibility criteria

We included full-text publications since 1990 with no language restrictions and including observational studies, qualitative studies, reports by professional bodies, systematic reviews and randomised controlled trials reporting on admissions of CYP (≤18 years) to any paediatric ward or adult general ward with a primary MH diagnosis. In studies where only average age was reported, studies were eligible if the average age of participants was ≤18 years. We excluded studies which exclusively reported on CYP presenting to the ED and those that reported admissions solely of participants aged >18 years.

### Search method for identification of studies

We searched PubMed, Embase, PsychINFO and Web of Science (1990 to April 2023). An additional search of Google Scholar was performed to identify reports which might contain unpublished data/additional studies. Search terms developed in conjunction with a clinical librarian were: (admission* OR admitted OR admittance OR hospitalized OR hospitalised OR treated OR inpatient* OR in patient* OR boarding OR boarders OR psychiatric boarders) and (paediatric ward* OR children* ward* OR pediatric ward*) and (mental health* OR psychiatric or psychological). Reference lists of selected articles were reviewed to identify additional studies.

### Study selection process

After duplicates were removed, two researchers (AVV, AS) independently reviewed titles and abstracts for inclusion. Differences were resolved by discussion with a third reviewer (LH). The same reviewers independently extracted information from selected studies to address the five review questions above.

### Quality assessment

The reviewers independently assessed included studies for quality. For qualitative studies, the Critical Appraisal Skills Programme (CASP) tool was used. This consists of 10 questions (scored as ‘yes’, ‘can’t tell’ or ‘no’) that address the rigour of the research methodology and the findings’ credibility. We then followed Fullen et al’s (17) proposal that if two-thirds scored ‘yes’, it was rated ‘high’, between four and six ‘yes’ was rated as ‘moderate’, and if over two-thirds were rated ‘no’, it was scored as ‘poor’ quality. For quantitative studies, the Appraisal tool for Cross-Sectional Studies (AXIS) was used. The AXIS tool aims to aid systematic interpretation of a study and to inform decisions about the quality of the study.

## RESULTS

### Description of included studies

Thirty-two studies met the inclusion criteria (Figure 1). The most common reasons for exclusion were full text unavailable, ED admissions only and irrelevance to our questions. Ten were US studies, seven were from the UK, six were from Australia, and the remaining were from Paraguay (n=1), Chile (n=1), France (n=1), Taiwan (n=2), Canada (n=1), the Republic of Ireland (n=2) and Germany (n=1). Detailed findings of the included studies are collated in Tables 1-4. Eighteen studies addressed trends and or numbers/proportions of admissions (3,4,6–8,18–30), 12 provided data about HCPs views/experiences (4,31–41), two provided data about CYP views/experiences (37,42) and four aimed at improving the care during admissions (6,40,43,44).

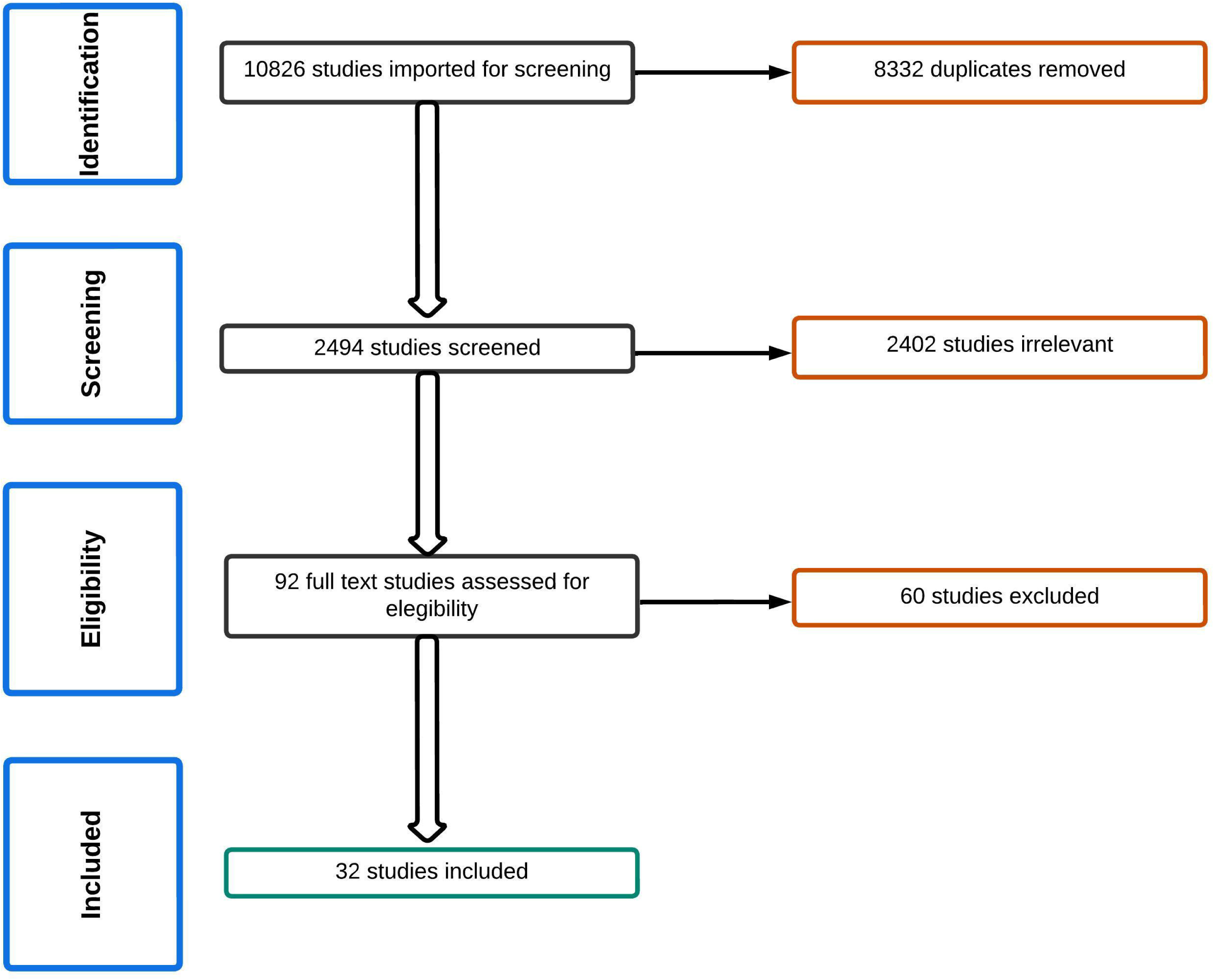

**Table 1.**
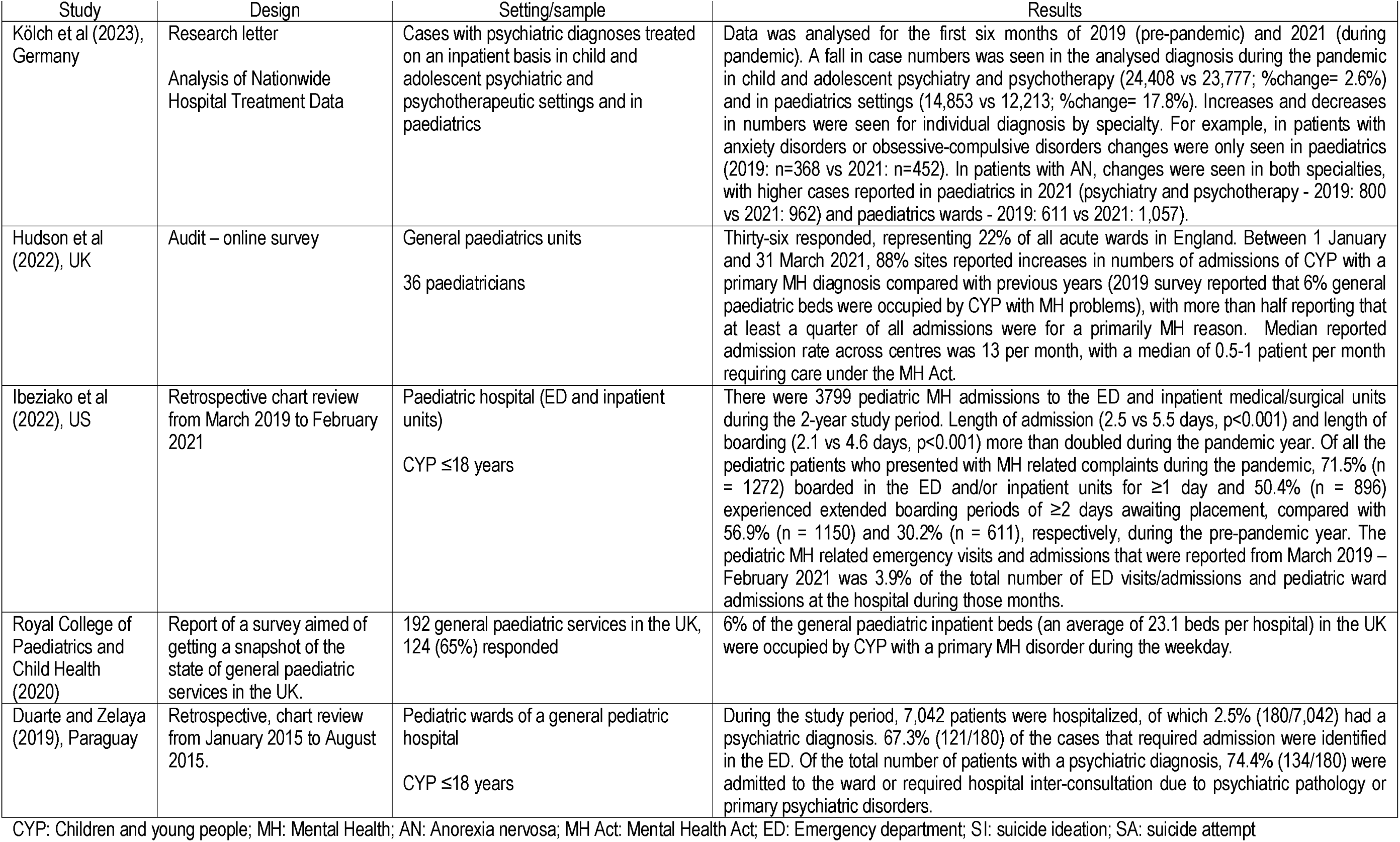
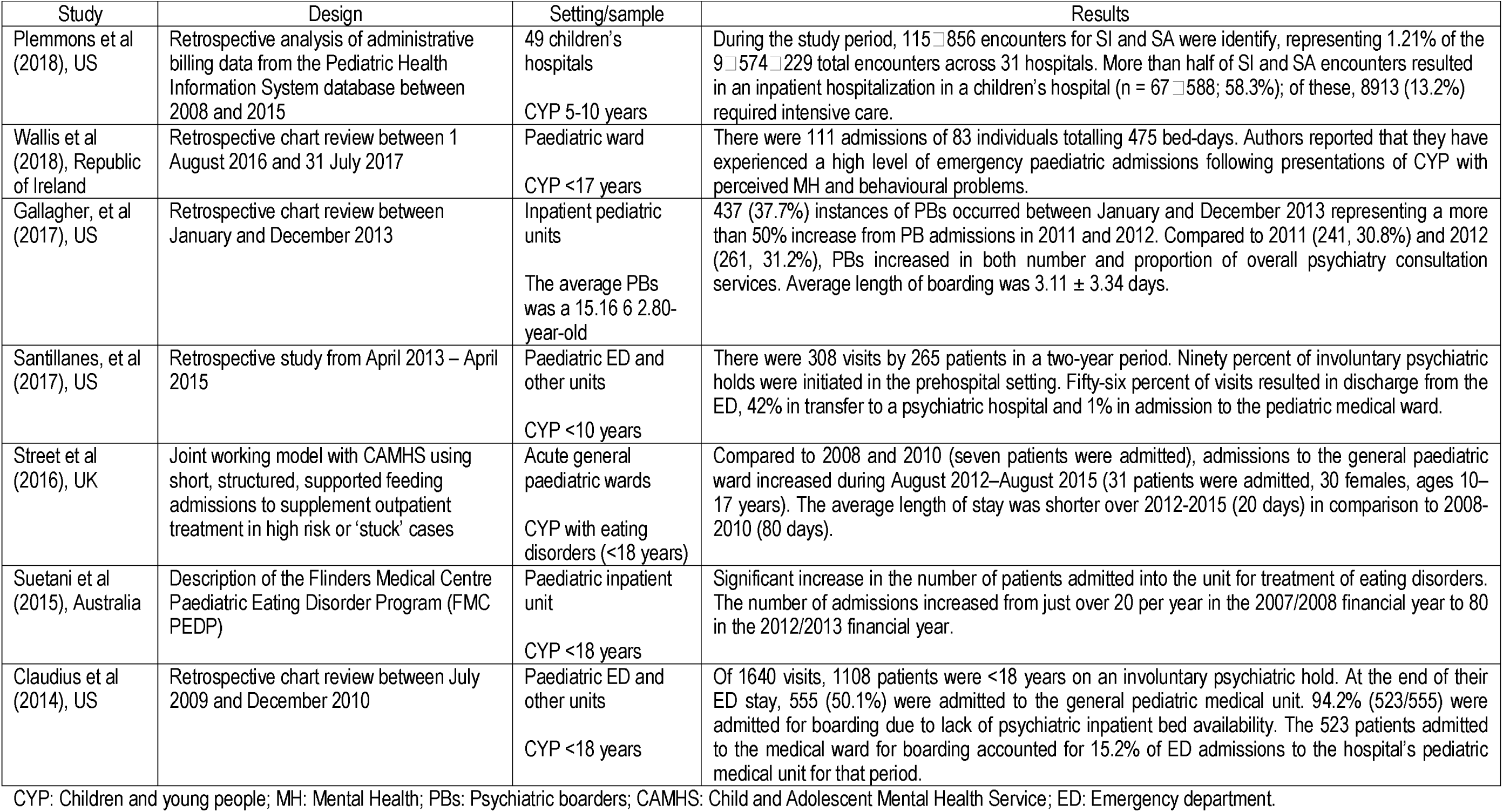
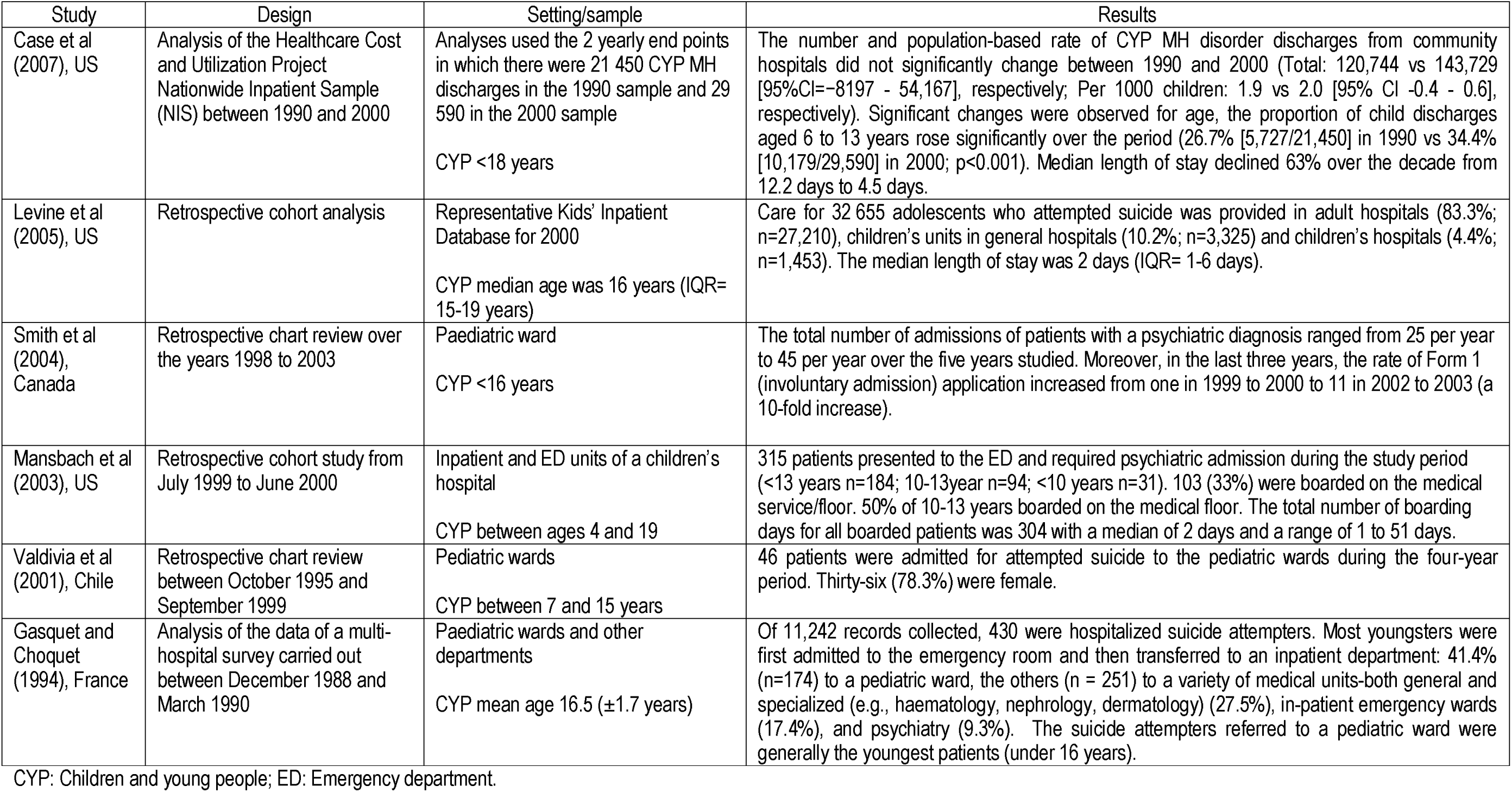
What is the trend in the number of admissions and the number/proportions of CYP admitted to paediatric wards or adult wards because of a MH diagnosis?

| Study | Design | Setting/sample | Results |
| --- | --- | --- | --- |
| Kölch et al (2023), Germany | Research letter<br>Analysis of Nationwide Hospital Treatment Data | Cases with psychiatric diagnoses treated on an inpatient basis in child and adolescent psychiatric and psychotherapeutic settings and in paediatrics | Data was analysed for the first six months of 2019 (pre-pandemic) and 2021 (during pandemic). A fall in case numbers was seen in the analysed diagnosis during the pandemic in child and adolescent psychiatry and psychotherapy (24,408 vs 23,777; %change= 2.6%) and in paediatrics settings (14,853 vs 12,213; %change= 17.8%). Increases and decreases in numbers were seen for individual diagnosis by specialty. For example, in patients with anxiety disorders or obsessive-compulsive disorders changes were only seen in paediatrics (2019: n=368 vs 2021: n=452). In patients with AN, changes were seen in both specialties, with higher cases reported in paediatrics in 2021 (psychiatry and psychotherapy - 2019: 800 vs 2021: 962) and paediatrics wards - 2019: 611 vs 2021: 1,057). |
| Hudson et al (2022), UK | Audit – online survey | General paediatrics units<br>36 paediatricians | Thirty-six responded, representing 22% of all acute wards in England. Between 1 January and 31 March 2021, 88% sites reported increases in numbers of admissions of CYP with a primary MH diagnosis compared with previous years (2019 survey reported that 6% general paediatric beds were occupied by CYP with MH problems), with more than half reporting that at least a quarter of all admissions were for a primarily MH reason. Median reported admission rate across centres was 13 per month, with a median of 0.5-1 patient per month requiring care under the MH Act. |
| Ibeziako et al (2022), US | Retrospective chart review from March 2019 to February 2021 | Paediatric hospital (ED and inpatient units)<br>CYP ≤18 years | There were 3799 pediatric MH admissions to the ED and inpatient medical/surgical units during the 2-year study period. Length of admission (2.5 vs 5.5 days, $p<0.001$) and length of boarding (2.1 vs 4.6 days, $p<0.001$) more than doubled during the pandemic year. Of all the pediatric patients who presented with MH related complaints during the pandemic, 71.5% (n = 1272) boarded in the ED and/or inpatient units for ≥1 day and 50.4% (n = 896) experienced extended boarding periods of ≥2 days awaiting placement, compared with 56.9% (n = 1150) and 30.2% (n = 611), respectively, during the pre-pandemic year. The pediatric MH related emergency visits and admissions that were reported from March 2019 – February 2021 was 3.9% of the total number of ED visits/admissions and pediatric ward admissions at the hospital during those months. |
| Royal College of Paediatrics and Child Health (2020) | Report of a survey aimed of getting a snapshot of the state of general paediatric services in the UK. | 192 general paediatric services in the UK, 124 (65%) responded | 6% of the general paediatric inpatient beds (an average of 23.1 beds per hospital) in the UK were occupied by CYP with a primary MH disorder during the weekday. |
| Duarte and Zelaya (2019), Paraguay | Retrospective, chart review from January 2015 to August 2015. | Pediatric wards of a general pediatric hospital<br>CYP ≤18 years | During the study period, 7,042 patients were hospitalized, of which 2.5% (180/7,042) had a psychiatric diagnosis. 67.3% (121/180) of the cases that required admission were identified in the ED. Of the total number of patients with a psychiatric diagnosis, 74.4% (134/180) were admitted to the ward or required hospital inter-consultation due to psychiatric pathology or primary psychiatric disorders. |

CYP: Children and young people; MH: Mental Health; AN: Anorexia nervosa; MH Act: Mental Health Act; ED: Emergency department; SI: suicide ideation; SA: suicide attempt

Table 1. Continued
| Study | Design | Setting/sample | Results |
| --- | --- | --- | --- |
| Plemmons et al (2018), US | Retrospective analysis of administrative billing data from the Pediatric Health Information System database between 2008 and 2015 | 49 children's hospitals<br>CYP 5-10 years | During the study period, 115 856 encounters for SI and SA were identify, representing 1.21% of the 9 574 229 total encounters across 31 hospitals. More than half of SI and SA encounters resulted in an inpatient hospitalization in a children's hospital (n = 67 588; 58.3%); of these, 8913 (13.2%) required intensive care. |
| Wallis et al (2018), Republic of Ireland | Retrospective chart review between 1 August 2016 and 31 July 2017 | Paediatric ward<br>CYP <17 years | There were 111 admissions of 83 individuals totalling 475 bed-days. Authors reported that they have experienced a high level of emergency paediatric admissions following presentations of CYP with perceived MH and behavioural problems. |
| Gallagher, et al (2017), US | Retrospective chart review between January and December 2013 | Inpatient pediatric units<br>The average PBs was a 15.16 6 2.80-year-old | 437 (37.7%) instances of PBs occurred between January and December 2013 representing a more than 50% increase from PB admissions in 2011 and 2012. Compared to 2011 (241, 30.8%) and 2012 (261, 31.2%), PBs increased in both number and proportion of overall psychiatry consultation services. Average length of boarding was 3.11 ± 3.34 days. |
| Santillanes, et al (2017), US | Retrospective study from April 2013 – April 2015 | Paediatric ED and other units<br>CYP <10 years | There were 308 visits by 265 patients in a two-year period. Ninety percent of involuntary psychiatric holds were initiated in the prehospital setting. Fifty-six percent of visits resulted in discharge from the ED, 42% in transfer to a psychiatric hospital and 1% in admission to the pediatric medical ward. |
| Street et al (2016), UK | Joint working model with CAMHS using short, structured, supported feeding admissions to supplement outpatient treatment in high risk or 'stuck' cases | Acute general paediatric wards<br>CYP with eating disorders (<18 years) | Compared to 2008 and 2010 (seven patients were admitted), admissions to the general paediatric ward increased during August 2012–August 2015 (31 patients were admitted, 30 females, ages 10–17 years). The average length of stay was shorter over 2012-2015 (20 days) in comparison to 2008-2010 (80 days). |
| Suetani et al (2015), Australia | Description of the Flinders Medical Centre Paediatric Eating Disorder Program (FMC PEDP) | Paediatric inpatient unit<br>CYP <18 years | Significant increase in the number of patients admitted into the unit for treatment of eating disorders. The number of admissions increased from just over 20 per year in the 2007/2008 financial year to 80 in the 2012/2013 financial year. |
| Claudius et al (2014), US | Retrospective chart review between July 2009 and December 2010 | Paediatric ED and other units<br>CYP <18 years | Of 1640 visits, 1108 patients were <18 years on an involuntary psychiatric hold. At the end of their ED stay, 555 (50.1%) were admitted to the general pediatric medical unit. 94.2% (523/555) were admitted for boarding due to lack of psychiatric inpatient bed availability. The 523 patients admitted to the medical ward for boarding accounted for 15.2% of ED admissions to the hospital's pediatric medical unit for that period. |

CYP: Children and young people; MH: Mental Health; PBs: Psychiatric boarders; CAMHS: Child and Adolescent Mental Health Service; ED: Emergency department.

Table 1 Continued
| Study | Design | Setting/sample | Results |
| --- | --- | --- | --- |
| Case et al (2007), US | Analysis of the Healthcare Cost and Utilization Project Nationwide Inpatient Sample (NIS) between 1990 and 2000 | Analyses used the 2 yearly end points in which there were 21 450 CYP MH discharges in the 1990 sample and 29 590 in the 2000 sample<br>CYP <18 years | The number and population-based rate of CYP MH disorder discharges from community hospitals did not significantly change between 1990 and 2000 (Total: 120,744 vs 143,729 [95%CI=-8197 - 54,167], respectively; Per 1000 children: 1.9 vs 2.0 [95% CI -0.4 - 0.6], respectively). Significant changes were observed for age, the proportion of child discharges aged 6 to 13 years rose significantly over the period (26.7% [5,727/21,450] in 1990 vs 34.4% [10,179/29,590] in 2000; $p<0.001$). Median length of stay declined 63% over the decade from 12.2 days to 4.5 days. |
| Levine et al (2005), US | Retrospective cohort analysis | Representative Kids' Inpatient Database for 2000<br>CYP median age was 16 years (IQR= 15-19 years) | Care for 32 655 adolescents who attempted suicide was provided in adult hospitals (83.3%; $n=27,210$), children's units in general hospitals (10.2%; $n=3,325$) and children's hospitals (4.4%; $n=1,453$). The median length of stay was 2 days (IQR= 1-6 days). |
| Smith et al (2004), Canada | Retrospective chart review over the years 1998 to 2003 | Paediatric ward<br>CYP <16 years | The total number of admissions of patients with a psychiatric diagnosis ranged from 25 per year to 45 per year over the five years studied. Moreover, in the last three years, the rate of Form 1 (involuntary admission) application increased from one in 1999 to 2000 to 11 in 2002 to 2003 (a 10-fold increase). |
| Mansbach et al (2003), US | Retrospective cohort study from July 1999 to June 2000 | Inpatient and ED units of a children's hospital<br>CYP between ages 4 and 19 | 315 patients presented to the ED and required psychiatric admission during the study period (<13 years $n=184$; 10-13year $n=94$; <10 years $n=31$). 103 (33%) were boarded on the medical service/floor. 50% of 10-13 years boarded on the medical floor. The total number of boarding days for all boarded patients was 304 with a median of 2 days and a range of 1 to 51 days. |
| Valdivia et al (2001), Chile | Retrospective chart review between October 1995 and September 1999 | Pediatric wards<br>CYP between 7 and 15 years | 46 patients were admitted for attempted suicide to the pediatric wards during the four-year period. Thirty-six (78.3%) were female. |
| Gasquet and Choquet (1994), France | Analysis of the data of a multi-hospital survey carried out between December 1988 and March 1990 | Paediatric wards and other departments<br>CYP mean age 16.5 ($\pm 1.7$ years) | Of 11,242 records collected, 430 were hospitalized suicide attempters. Most youngsters were first admitted to the emergency room and then transferred to an inpatient department: 41.4% ($n=174$) to a pediatric ward, the others ($n = 251$) to a variety of medical units-both general and specialized (e.g., haematology, nephrology, dermatology) (27.5%), in-patient emergency wards (17.4%), and psychiatry (9.3%). The suicide attempters referred to a pediatric ward were generally the youngest patients (under 16 years). |

CYP: Children and young people; ED: Emergency department.

### Trends/number/proportions of admissions of CYP

We found 18 studies reporting numbers and proportions of primary MH admissions of CYP ≤18 years to paediatric settings (Table 1). Nine used a retrospective chart review design for reporting admissions to single hospitals (7,8,18,21–23,26,28,29). Ibeziako et al (7) reported 3799 paediatric MH admissions to the ED and inpatient units at a paediatric hospital from March 2019 – February 2021. Duarte and Zelaya (26) reported 180 admissions of patients with psychiatric diagnoses (January-August 2015); 74.4% required admission to the paediatric ward or hospital inter-consultation because of psychiatric pathology or primary psychiatric disorders. Wallis (28) reported 111 emergency admissions (83 patients) of CYP with MH needs to the paediatric ward between August 2017 and July 2017. Gallagher et al (8) reported 437 PBs admissions on inpatient paediatric units between January and December 2013. Santillanes et al (29) reported 308 visits (265 patients on involuntary psychiatric hold) from April 2013 to April 2015; 1% of visits resulted in admissions to the paediatric ward. Claudius et al (18) reported 1108 patients on an involuntary psychiatric hold between July 2009 and December 2010; 50.1% were admitted to the general paediatric medical unit. Smith et al (21) reported that yearly admissions to the paediatric unit of patients with a psychiatric diagnosis ranged from 25 per year to 45 per year over the five years studied (1998 to 2003). Mansbach et al (22) reported 315 paediatric admissions at inpatient and ED units from July 1999 to June 2000; 33% were boarded on the medical/service floor. Valdivia et al (23) reported 46 patients admitted for suicide attempted (SA) at a paediatric ward between October 1995 and September 1999.

Four studies analysed large databases that included the reporting of MH admissions and discharges (19,20,25,27). Using the Paediatric Health Information database Plemmons et al (27) identified, between 2008 and 2015, 115,856 SA and suicidal ideations (SI) encounters across 31 hospitals of which 67,588 resulted in an inpatient hospitalization in a children’s hospital. Using the representative Kids’ database for 2000, Levine et al (20) reported that care for SA patients (n= 32,655) was provided in adult hospitals (83.3%), children’s units (10.2%) and children’s hospitals (4.4%). Using the Nationwide Inpatient Sample (NIS), Case et al (19) analysed data between 1900 and 2000 (n≈1000 hospitals) reporting non-significant changes in CYP MH disorders discharges from community hospitals (Per 1000 children: 1.9 vs 2.0 [95% CI -0.4 - 0.6], respectively). However, CYP discharges ages 6-13 years rose significantly (26.7% [5,727/21,450] in 1990 vs 34.4% [10,179/29,590] in 2000; p<0.001). Finally, Kölch et al (25) analysed data for MH admissions in CYP from Germany, comparing the first six months of 2019 (pre-pandemic) and of 2021 (during the pandemic). They found no change in the number of admissions to specialist MH inpatient care for CYP with anxiety disorders or obsessive-compulsive disorders between time points. However, there was increase in patients with anorexia nervosa (AN) to both general paediatric wards and specialist MH inpatient setting, with a higher burden of cases reported in paediatrics wards - 2019: 611 vs 2021: 1,057).

Three studies reported data from surveys. Hudson et al (4) surveyed paediatricians working in acute paediatric services in England and received responses from 22% of all acute wards in England; they found that 88% of respondents reported increases in MH admissions between January and March 2021 (4). Gasquet and Choquet (24) reported 430/11,242 SA records between December 1988 and March 1990 among 164 hospitals; 174/430 patients were admitted to the paediatric wards (24). Royal College of Paediatrics and Child Health surveyed all general paediatric services in the UK in 2019 and found that across sites 6% of the general paediatric inpatient beds in the UK were occupied by CYP with a primary MH disorder (3). Finally, two studies that describe the development/implementation of programs for patients with eating disorders (EDs) reported, as part of this description, the number of admissions. Street et al (9) reported that from August 2012–August 2015, 31 patients with EDs were admitted to the general paediatric ward in Exeter. Compared to admissions between 2008 and 2010 (seven admissions), admissions increased. Suetani et al (30) reported an increase in the number of patients admitted into the paediatric inpatient unit for treatment of EDs at the Flinders Medical Centre in Australia. From over 20 per year in 2007/2008 to 80 in 2012/2013.

### HCPs’ experiences

Twelve papers reported experiences of HCPs (Table 2). Six were qualitative (semi-structured or in-depth interviews and focus groups) (31,33,35–37,39) and two mixed-method (38,41). These studies used a range of epistemological perspectives (grounded theory, content analysis, thematic analysis and phenomenology) for data analysis. Four other observational studies used a questionnaire to survey HCPs caring for CYP during admissions (4,32,40), with one applying thematic content analysis using data derived from open-ended questions (34). Eight studies provided evidence suggesting that a concern of HCPs was lack of skills/knowledge and confidence to care for CYP admitted in acute paediatric wards (4,31,32,34,36,39–41). Four studies also reported HCPs’ concerns about the appropriateness of paediatric ward environments for the treatment of this group of patients. Commonly, HCPs reported difficulty in focusing on patients with MH problems in the acute ward due to the busy and complex make-up of patients across wards, and stressed the need for separate units/rooms to treat this group (32,35,38,39). Other reported experiences was a lack of support from MH professionals (4,40), feeling frustration because of the lack of knowledge/time/resources while caring for this group (33,40,41) and the difficulty of establishing therapeutic relationships (31,35,41). HCPs however reported their desire for more knowledge about MH resources and how to safely allocate and plan care for them (36) and also positive impacts of training applied to experience caring for CYP with mental health problems to enhance competence/confidence (32,34).

**Table 2.** What are the reported experiences of clinical staff on paediatric wards (or adult general wards) during the admissions of CYP admitted because of a primary MH diagnosis?

| Study | Design | Setting/sample | Results |
| --- | --- | --- | --- |
| Chang et al (2023), Taiwan | Qualitative study - semi-structured interviews<br>Content analysis approach | General paediatric ward<br>16 HCPs, including 10 nurses, 3 dieticians, and 3 paediatric gastroenterologists | 1. Building a trusting relationship first: HCPs cannot easily establish a therapeutic relationship with a patient at the first encounter. Patients are highly defensive and reluctant to express her thoughts and feelings.<br>2. The key to treatment success: The most difficult aspect of the treatment and care of adolescents with anorexia is whether the patient understands that they have an illness.<br>3. Consistency of team treatment goals: The nurse and the patient set a weight goal together, and upon achieving the goal, the patient will be allowed outside or discharged from the hospital.<br>4. Empowerment with knowledge about anorexia: Participants described the lack of knowledge of HCPs, especially nurses and dieticians, in the care of anorexia and the expectation of continuing education related to anorexia.<br>5. Using different interaction strategies: Some participants would use coercive methods while others use gentleness or physical comfort. |
| Hudson et al (2022), UK | Cross-sectional – online survey | General paediatrics units<br>36 paediatricians | In free-text responses, paediatricians reported a lack of MH support and insufficient skills and training, for example restraint practices. |
| Lakeman and McIntosh (2018), Australia | Quantitative and qualitative data – online survey with open-ended questions<br>Thematic content analysis using data derived from open-ended questions | Emergency department, medical, paediatric wards and MH services of the Cairns and Hinterland Hospital and Health Service (CHHS)<br>136 clinicians working with patients with EDs | 73% reported little or no confidence working with EDs. Those who reported 70 or more hours of training were 2.7 times more likely to report feeling competent and confident (OR = 2.7, CI = 95%, $p < 0.05$). |
| Wu and Chen (2021), Taiwan | Qualitative exploratory study – semi-structured interview<br>Content analysis approach | General paediatric ward at a Children's Hospital<br>10 nurses | 1. Struggling to develop therapeutic relationships: Patients with AN tend to be very defensive, so it is not easy for nurses and patients to establish a therapeutic relationship.<br>2. Selective focusing: Due to the nature of the acute ward, nursing staff often need to take care of multiple patients simultaneously, which means insufficient time interacting with patients and lack of positive feelings in the AN patient care.<br>3. Difficulty changing minds: Refers to the fact that patients with AN usually lack a sense of illness. They are involuntarily hospitalized, so they passively cooperate with medical treatment. |

CYP: Children and young people; MH: Mental Health; HCPs: Health care professionals; EDs: Eating disorders; AN: Anorexia.

Table 2 Continued
| Study | Design | Setting/sample | Results |
| --- | --- | --- | --- |
| Hampton et al (2015), US | Qualitative study – focus groups<br>Grounded theory method | Urban paediatric clinics<br>26 paediatric residents | 1. Capabilities: Residents expressed uncertainty regarding knowledge and skills about MH care.<br>2. Comfort: Residents predominantly expressed discomfort with the provision of MH care.<br>3. Organizational capacity: Time limitations and continuity of care were specifically mentioned as barriers within their clinic.<br>4. Coping: They coped by reducing their scope of medical practice by triaging and referring MH care rather than accepting more responsibility.<br>5. Education: Residents desired more knowledge of what MH resources exist, how to appropriately allocate them, the processes for making referrals, and the strategies for managing patients with specialists. |
| Ramjan and Gill (2012), Australia | Qualitative study – semi-structured interview<br>Thematic analysis<br>This study examined an inpatient behavioural program for adolescents with AN | One adolescent ward in an acute care pediatric setting<br>10 paediatric nurses | In general, nurses believed the program's intentions were "honourable" and that they had a duty to follow the program. However, having the role of "prison warden" diffculted the development of therapeutic relationships. Moreover, caring for adolescent patients in this program became "very routine" and "monotonous" for most nurses. Nearly all saw themselves go into "autopilot" on the ward because they "[knew] the routine inside out." |
| Buckley (2010), Republic of Ireland | Exploratory mixed methods approach (descriptive statistics and qualitative findings) - Content analysis<br>'Questionnaire on Nurse's Experiences of Nursing Young People with MH Problems in the Paediatric Ward Setting' (adapted from The Adolescent Mental Health Nursing Questionnaire). | General paediatric wards<br>39 registered staff nurses | Most nurses (66.6%) were not satisfied with their ability to care for CYP with MH conditions. A total of 76.7% of nurses highlighted that the most difficult problem is nursing a CYP with a MH condition on an acute medical/surgical unit. Overall, 67% of nurses were dissatisfied with having to nurse this group on a general paediatric ward. 86.6% stated that this group should be cared for in separate units. A total of 15.3% of this 86.6% response rate indicated that this group required single rooms on the general paediatric wards. |
| Happell et al (2009), Australia | Participatory action research – Focus groups<br>Thematic analysis | Paediatric unit of a rural general hospital<br>A purposive convenience sample of all paediatric nursing staff (n = 20; of 24 nurses). | MH care was identified as particularly problematic because of the absence of clear clinical pathways. A lack of understanding of general nurses' role in the management of CYP admitted to the paediatric unit with an acute mental illness, meant participants' confidence in caring for such patients was affected. Participants worried that the unit was not always a place of safety, given the occasionally unpredictable nature of adolescents, particularly those with a MH issue. Participants were concerned that their lack of training and experience with patients with MH issues was detrimental to the delivery of optimal patient care. |

CYP: Children and young people; MH: Mental Health; AN: Anorexia.

Table 2 Continued
| Study | Design | Setting/sample | Results |
| --- | --- | --- | --- |
| Watson (2006), UK | Survey to identify nurse's concerns and attitudes | General wards - Birmingham Children's Hospital National Health Service (NHS) Trust<br>90 nurses | Sixty-four per cent of those who responded said they nursed CYP with MH issues in their clinical area, and 79% stated that they did not feel experienced in meeting the needs of this group. Moreover, 67% reported little or no support from MH professionals. Nurses appeared to have little knowledge of CAMHS provision. Most (84%) agreed that this is what frustrated them indicating a need to raise awareness about CAMHS structure, input, roles and assessment procedures throughout the trust. This lack of awareness may explain why most respondents felt the trust does not do enough for this group. |
| Anderson et al (2003), UK | Quantitative and qualitative methods (semi-structured interviews), and a contemporary grounded theory approach to analysis | A&E, paediatric medicine and child and adolescent MH services<br>45 HCPs | Experiences of frustration in practice: Not having enough time and resources to enhance their relationships with CYP who had engaged in suicidal behaviour.<br>Strategies for relating to CYP: Nurses working in paediatric medicine and A&E came a realisation that they were less able to offer specific skills (i.e., competency in communicating with this patient group). |
| Ramritu et al (2002), Australia | Questionnaire (Adolescent Mental Health Nursing Questionnaire [AMHNQ]) to survey registered nurses | Paediatric tertiary referral hospital (one acute medical ward and one combined acute medical/surgical ward)<br>30 generalist nurses | 1. Experience in care provision: 57% of participants felt confident in assessing changes in the adolescents' MH and 30% did not feel confident. There was a significant association between years of MH experience and confidence in assessing this group ($p<0.02$) and between the years of MH experience and the usefulness of in-service education provided by MH nurses ($p<0.03$).<br>2. Satisfaction with ability to care for adolescents: 40% were satisfied with their ability to care for this group.<br>3. Challenges encountered in care: 90% stated that they encounter problems in caring for this group. The most frequently identified problem was nursing adolescents in an acute medical ward. |
| King and Turner (2000), Australia | Qualitative research - Audio-taped in-depth interviews<br>Phenomenology | Paediatric wards of general hospital<br>Five registered nurses without psychiatric nursing or MH qualifications | Succinct statement of the phenomenon (6 emergent themes were interwoven): Caring for adolescent females with anorexia nervosa was a journey of frustration. A turmoil of emotions was experienced, which inevitably eroded their resolve of maintaining core nursing values. The feeling of failure and loss of faith was the nadir of despair in the experience. This negative self-image impelled them to change their focus and redirect their efforts to understand the reality of the predicament of the anorexics. This became the pivot for altering attitudes and building resolutions that enabled them to care for their patients. |

CYP: Children and young people; MH: Mental Health; HCPs: Health care professionals; CAMHS: Child and Adolescent Mental Health Service; A&E: Accident and emergency.

### CYPs’ experiences

We found two qualitative studies examining CYP experiences during admissions (37,42) (Table 3). Worsley et al (42) explored the experiences of adolescents during boarding hospitalization following SI or SA (n=27). Participants expressed appreciation for compassionate clinicians and for information about what to expect during their hospital stay. Ramjan and Gil (37) interviewed 10 adolescents with anorexia (AN) admitted to the acute care paediatric setting within an inpatient behavioural program. One participant described her first admission as a “terrible, traumatic” experience. Others recalled emotions, including fear, anger, depression, and confusion.

**Table 3.** What are the reported experiences of CYP and their families during admissions to paediatric wards (or adult general wards) because of a primary MH diagnosis?

| Study | Design | Setting/sample | Results |
| --- | --- | --- | --- |
| Worsley et al (2019), US | Qualitative design using semi-structured interviews<br>Conventional content analysis | Inpatient unit of a children's hospital<br>Convenience sample of 27 CYP (9 – 18 years) hospitalized for SI or SA while they were awaiting transfer to an inpatient psychiatric facility | Specifically, adolescents felt more secure when clinicians described the processes of the emergency department visit, pediatric hospitalization, and inpatient psychiatric hospitalization to them; this helped them feel less stressed about the current hospitalization and the plan for an inpatient psychiatric hospitalization. Adolescents expressed interest in receiving several types of information about psychiatric hospitalization: food, visitation policies, length of stay, entertainment, daily activities and schedules, location, clinicians providing treatment, types of therapy provided, and the physical structure and layout of inpatient psychiatric units. Many participants described feelings of stress, anxiety, and embarrassment when they were asked repeatedly by different clinicians to explain their health history and reason for hospitalization.<br>Many adolescents compared their current hospitalization with previous medical hospital experiences. For some patients, being in a medical hospital felt familiar and comfortable. For other patients, fears related to previous medical experiences emerged; several patients worried about the possibility of painful treatment. |
| Ramjan and Gil (2012), Australia | Qualitative, naturalistic design, using in-depth, face-to-face, semi structured interviews<br>Thematic analysis<br>This study examined an inpatient behavioural program for adolescents with AN | One adolescent ward in an acute care pediatric setting<br>10 adolescent patients | Adolescents entered the system in one of two ways. Either they were taken to the emergency department by a concerned family member, or they were attending a clinic appointment when the decision was made to admit them.<br>One participant described her first admission as a “terrible, traumatic” experience. Others recalled many emotions, including fear, anger, depression, and confusion, about why they were being admitted. Another participant “never thought that someone could come into hospital for that kind of condition,” and it made her think “I shouldn't be in here.” Another thought she was “en route for a holiday” when her family suddenly admitted her for treatment. As she recalls the day: “I didn't even know we were stopping at the hospital. We were stopping in for counselling or something. I didn't know.... Then I found out straightaway that I was being admitted and my parents had to leave within ... half an hour of dropping me off.” |

CYP: Children and young people; MH: Mental Health; SI: suicide ideation; SA: suicide attempt; AN: Anorexia

**Table 4.** Is there evidence of interventions or quality improvement projects aimed at improving the care of CYP and families during admissions to paediatric wards (or adult wards) because of a primary MH diagnosis?

| Study | Design | Setting/sample | Results |
| --- | --- | --- | --- |
| Todd et al (2023), UK | An audit of care was carried out followed by a MH teaching week with clinical staff to improve quality of care and staff confidence when working with CYP with MH issues. | Acute paediatric ward of a general paediatric department in a London district general hospital<br>15 responses prior teaching week<br>9 responses after teaching week | Staff confidence prior to the multidisciplinary teaching week showed that no doctors felt 'very confident' when reviewing CAMHS patients, with 60% feeling 'somewhat confident' and 19% feeling 'not confident' (n=15). After the teaching week, 89% reported that the teaching week had improved their confidence in managing MH presentations and 100% said that more teaching on this subject would be beneficial. The MH simulation scenario on taking a history from a suicidal adolescent was thought to be the most useful session, followed by teaching from the CAMHS team on the use of rapid tranquilisation in paediatrics and fellow paediatric trainees. However, there were no sustained improvements in the care of MH patients when comparing the Audit from 1 March 2021 with the post-intervention audit from January 2022. |
| Bolland et al (2017), UK | The authors carried out an interactive workshop to promote CHCPs' communication skills with CYP who have MH needs | General children's wards<br>34 generalist CHCPs | The workshop was divided into two sessions:<br>Session 1: Delivered by Redthread, a youth work charity. The session focused on communicating with CYP and engaging with them. Participants' perceptions were challenged by using visual exercises and asking them what they saw to highlight the differences in perceptions and the need to be aware of personal biases.<br>Session 2: Delivered by an expert in child and adolescent MH who works in the CAMHS. Communication with CYP with MH needs, with the focus on self-harm and EDs, was explored. Participants were introduced to the PATHOS screening instruments for overdose. This enabled the participants to build on the communication skills developed during session 1.<br>Participants were asked to complete an evaluation of the workshop. All completed the evaluation and reported that the workshop provided them with tools and strategies to try in practice. Their confidence increased because of the workshop, and they had more positive attitudes toward CYP. In terms of long-term benefits, six weeks after the workshop, five CHCPs provided a reflexion report. There was evidence of improved communication skills and participants felt more confident when communicating with CYP. |

CYP: Children and young people; MH: Mental Health; CAMHS: Child and Adolescent Mental Health Service; CHCPs: children's healthcare professionals; EDs: eating disorders.

Table 4 Continued
| Study | Design | Setting/sample | Results |
| --- | --- | --- | --- |
| Street et al (2016), UK | Development of a joint working model with CAMHS using short, structured, supported feeding admissions to supplement outpatient treatment in high risk or 'stuck' cases | Acute general paediatric wards<br>31 patients admitted ages 10-17 years | Local paediatric wards successfully managed most young people in the community avoiding lengthy, expensive, specialist CAMHS-ED admissions (tier 4). Local ward admissions are easier to manage and the change in ward admissions has created a more positive attitude among staff towards CYP. Key to success has been communication and joint working between professionals, and the removal of the artificial divide between physical and MH, and medical and CAMHS teams. |
| Watson (2006), UK | Cross-sectional – questionnaire<br>Based on the findings reported in Table 2 (see Watson 2006), a project was initiated to improve nursing liaison with CAMHS nurses providing support and advice to general children's nurses. | General wards - Birmingham Children's Hospital National Health Service (NHS) Trust<br>90 nurses | Once the liaison service was initiated, the biggest component quickly became teaching and education. A one-day study event that soon extended to a two-day programme enabled paediatric nursing colleagues to become better informed on the holistic aspects of MH care. The most significant outcome of the programme was increased awareness of MH issues and the informal discussions generated within paediatric environments. This culminated in the formation of a MH interest group by children's nurses in the trust. Informal feedback indicated that nurses were liberated by being able to contact their CAMHS colleagues for telephone advice and guidance; they were able to question their current or traditional practices. Armed with evidence-based material, nurses were more confident in challenging approaches and attitudes of paediatricians and other disciplines as they established new working practices and methods for care delivery. |

CYP: Children and young people; MH: Mental Health; CAMHS: Child and Adolescent Mental Health Service; CAMHS-ED: Child and Adolescent Mental Health Service Eating Disorders inpatient unit.

### Improving the care of CYP and their families/carers during admissions

We found four studies aimed at improving the care of CYP during admissions (6,40,43,44) (Table 4). Todd et al (43) carried out a MH teaching week with HCPs to improve quality of care/confidence when working with this group. Overall, after the teaching session, 89% per cent reported improvement in their confidence in managing MH presentations in paediatrics. However, there were no sustained improvements in the care of MH patients when comparing the audit from March 2021 (pre-teaching week) with the post-teaching week audit (January 2022). Bolland et al (44) carried out an interactive workshop to promote HCPs’ communication skills with CYP with MH needs. Participants (n=34) completed an evaluation of the session and reported that the workshop provided them with tools/strategies to try in practice. Six weeks after the workshop, there was evidence of improved communication skills and participants felt more confident when communicating with CYP. Street et al (9) developed a joint working model with Child and Adolescent Mental Health Services (CAMHS) to avoid specialist CAMHS-eating disorders inpatient unit admissions. They reported positive impacts provided by communication and joint working between professionals, in particular between physical health and MH professionals. Watson et al (37) reported on a project to improve paediatric nursing liaison with CAMHS nurses providing support/advice to paediatric nurses. A two-day programme was carried out which aimed to enable nurses to become better informed on the holistic aspects of MH care. Feedback indicated that nurses felt able to contact CAMHS colleagues for advice/guidance. Nurses were more confident in challenging approaches/attitudes of paediatricians/other disciplines as they established new working practices/methods for care.

### Quality assessment

We assessed nine studies using the CASP tool (Table S1). Six studies were rated high quality (31,35,36,39,41,42), which represents 67% of the total studies assessed (n=9), two (33,37) moderate quality (22%) and only one (40) low quality (11%). We assessed fifteen studies using the AXIS scale (Table S2). In 11 studies (73%) it was unclear what methods were used to determine the sample size (7,8,18,21–25,28,29,32). Only one study (7%) provided clear information about the measurements undertaken to address non-response (22), and none reported clear information about concerns around non-response bias. Five studies (33%) did not provide clear methods to determine statistical significance or precision estimates (8,18,21,24,28) and 10 (67%) did not disclose if funding sources or conflicts of interest might affect authors’ interpretation of the results (19–24,26,28,32).

We were unable to assess two mixed methods because of the lack of a clear mixed-method question/objectives (38) and insufficient information on the qualitative methods to address the data collection (34) (see screening questions of the Mixed Methods Appraisal Tool 2018- http://mixedmethodsappraisaltoolpublic.pbworks.com/). One cross-sectional study was not assessed due insufficient information on the methodology (3). Moreover, we did not find an appropriate tool that allowed us to assess studies that focused on describing the implementation/description of workshops, teaching weeks, working models/programs and clinical audits (4,6,30,43,44).

## DISCUSSION

To our knowledge, this is the first systematic review on CYP admissions to paediatric wards with a primary MH indication. We found a range of studies reporting on numbers of such admissions indicating that these admissions are common across a range of countries, however only a small number of studies addressed trends over time. Those that did suggested increased numbers over time, especially since the pandemic. Reasons cited for increased admissions in those papers included lack of joint working between paediatric medical and MH services (6), unavailability of inpatient psychiatric placements (7,8,22), shortage of paediatric liaison psychiatry services (28) and the increasing prevalence of MH conditions in CYP such as suicide ideation or attempt and depressive disorders (19,27). We also found evidence of HCP working on paediatric wards of concerns about skill sets to manage CYP with MH presentations, and from some questioning the appropriateness of the acute ward for this care. Specific concerns included a lack of guidelines or standards for delivering care in this acute setting (28), lack of knowledge about what MH resources exist and how to allocate them (36), little knowledge of CAMHS provision (40), lack of separate units in the ward to care/treat this group (32,35,38,39) and not being able to offer specific skills, such as competency in communicating with this group (41) and restraint practices (4). Available evidence of CYP experiences was very limited and we found no studies on families/carers experiences. A main finding from CYP experience was a need for clear communication and compassionate clinicians caring for them. Finally, we found a limited number of studies reporting efforts to improve care of CYP during admission. These were all service evaluation papers rather than trials, limiting the quality of evidence provided, but they highlighted the importance of co-working and training to improve competencies and confidences, albeit with a need for repetition of training over tome to maintain these. We found no published evidence of specific risk factors for adverse care for CYP and families/carers during admissions.

Our review therefore provides important information for care of CYP admitted to general paediatric wards as well as key areas of need for further research. Better training and support for staff, as well as clear communication to CYP through there admission are important. Training opportunities may need to be repeated to ensure sustained impact. Joint working, between professionals with physical health and MH expertise also appears important, fraught as this is with availability, and calls for joint training across professions for this domain. Whilst several papers have reported absolute numbers, there is a clear need for bigger studies utilizing nationally available data on trends of admissions to better inform and plan care and workforce needs at both a local and national level. Numbers of studies examining CYP and carer experience and needs is lacking and requires more study. Lastly there is a clear need for the development of interventions to improve experience and quality of care for CYP admitted to paediatric wards and where possible these interventions should be tested and reported with better quality methodology such as trials. Given CYP’s experiences, such studies should utilize the input of CYP and carers in co-design.

### Strengths and limitations

We conducted a broad search across a range of important questions on this topic using five databases, and with independent screening of study eligibility. That said, despite finding sufficient suggestive evidence for clinical and research recommendations, we found few relevant studies, generally with small sample sizes and of limited in quality in relation to the questions we were asking. Although we carried out a Google Scholar search to identify unpublished data and snowballed references, we know that paediatric centres frequently have unpublished audits and service evaluations which we will have missed.

In summary, for services to be delivered effectively, for CYPs and their families/carers to feel supported and HCPs to feel confident, we need to strengthen the evidence base, but meanwhile to facilitate more robustly evaluated integrated physical and MH pathways of care, better (and regular) training and communication to CYP. These admissions are common and appropriate and safe care requires a significant increase in the amount and quality of research to provide this.

## Supporting information

Supplementary file_Quality assessment

Supplmentary file_PROSPERO

PRISMA_checklist

## Acknowledgements

We would like to thank Heather Chesters, Deputy Librarian at UCL Great Ormond Street Institute of Child Health Library, for supporting this work.

## Contributors

LH, FH, RV conceived the study. LH and AVV designed the search strategy. AVV and AS carried out the literature searches. AVV and AS screened the titles, abstracts and full texts and carried out the data extraction and quality assessment. AVV wrote the first draft and LH and HR were involved in the interpretation of data and provided valuable contribution towards reviewing, editing and completion of the final draft. All authors had access to all the data in the study and the responsibility for the decision to submit for publication.

## Data availability

All data relevant to the review are included in the article or uploaded as supplementary information.

## REREFENCES

1. World Health Organization. Adolescent mental health [Internet]. 2021. Available from: https://www.who.int/news-room/fact-sheets/detail/adolescent-mental-health

2. Clisu DA, Layther I, Dover D, Viner RM, Read T, Cheesman D, et al. Alternatives to mental health admissions for children and adolescents experiencing mental health crises: A systematic review of the literature. Clin Child Psychol Psychiatry [Internet]. 2022;27(1):35–60. Available from: 10.1177/13591045211044743

3. Royal College of Paediatrics and Child Health. A snapshot of general paediatric services and workforce in the UK [Internet]. 2020. Available from: https://www.rcpch.ac.uk/sites/default/files/2020-09/a_snapshot_of_general_paediatric_services_and_workforce_in_the_uk_1.4.pdf

4. Hudson LD, Chapman S, Street KN, Nicholls D, Roland D, Dubicka B, et al. Increased admissions to paediatric wards with a primary mental health diagnosis: results of a survey of a network of eating disorder paediatricians in England. Arch Dis Child [Internet]. 2022 Mar 1;107(3):309 LP – 310. Available from: http://adc.bmj.com/content/107/3/309.abstract

5. Shekunov J, Lewis CP, Vande Voort JL, Bostwick JM, Romanowicz M. Clinical Characteristics, Outcomes, Disposition, and Acute Care of Children and Adolescents Treated for Acetaminophen Toxicity. Psychiatr Serv [Internet]. 2021;72(7):758–65. Available from: 10.1176/appi.ps.202000081

6. Street K, Costelloe S, Wootton M, Upton S, Brough J. Structured, supported feeding admissions for restrictive eating disorders on paediatric wards. Arch Dis Child [Internet]. 2016 Sep 1;101(9):836 LP – 838. Available from: http://adc.bmj.com/content/101/9/836.abstract

7. Ibeziako P, Kaufman K, Scheer KN, Sideridis G. Pediatric Mental Health Presentations and Boarding: First Year of the COVID-19 Pandemic. Hosp Pediatr [Internet]. 2022 Aug 1;12(9):751–60. Available from: 10.1542/hpeds.2022-006555

8. Gallagher KAS, Bujoreanu IS, Cheung P, Choi C, Golden S, Brodziak K, et al. Psychiatric Boarding in the Pediatric Inpatient Medical Setting: A Retrospective Analysis. Hosp Pediatr [Internet]. 2017 Aug 1;7(8):444–50. Available from: 10.1542/hpeds.2017-0005

9. Chapman S, Hudson LD, Street KN. Restrictive eating disorders in children and young people: the role of the paediatrician and paediatric ward. Arch Dis Child [Internet]. 2022; Available from: https://adc.bmj.com/content/early/2022/02/06/archdischild-2021-322745

10. Morris J, Fisher E. Growing problems, in depth: The impact of Covid-19 on health care for children and young people in England [Internet]. Nuffieldtrust. 2022. Available from: https://www.nuffieldtrust.org.uk/resource/growing-problems-in-detail-covid-19-s-impact-on-health-care-for-children-and-young-people-in-england

11. Otis M, Barber S, Amet M, Nicholls D. Models of integrated care for young people experiencing medical emergencies related to mental illness: a realist systematic review. Eur Child Adolesc Psychiatry [Internet]. 2022 Sep 24; Available from: https://link.springer.com/10.1007/s00787-022-02085-5

12. Feng JY, Toomey SL, Zaslavsky AM, Nakamura MM, Schuster MA. Readmission After Pediatric Mental Health Admissions. Pediatrics [Internet]. 2017 Dec 1;140(6):e20171571. Available from: 10.1542/peds.2017-1571

13. Royal College of Paediatrics and Child, Royal College of Emergency Medicine, Royal College of Psychiatrists. Meeting the mental health needs of children and young people in acute hospitals: these patients are all our patients [Internet]. 2021. Available from: https://www.rcpch.ac.uk/resources/mental-health-needs-children-young-people-acute-hospitals

14. Cadorna G, Vera San Juan N, Staples H, Johnson S, Appleton R. Review: Systematic review and metasynthesis of qualitative literature on young people’s experiences of going to A&E/emergency departments for mental health support. Child Adolesc Ment Health [Internet]. 2023 Oct 12; Available from: https://acamh.onlinelibrary.wiley.com/doi/10.1111/camh.12683

15. Newton AS, Hartling L, Soleimani A, Kirkland S, Dyson MP, Cappelli M. A systematic review of management strategies for children’s mental health care in the emergency department: update on evidence and recommendations for clinical practice and research. Emerg Med J [Internet]. 2017 Jun;34(6):376–84. Available from: https://emj.bmj.com/lookup/doi/10.1136/emermed-2016-205939

16. Hoge MA, Vanderploeg J, Paris M, Lang JM, Olezeski C. Emergency Department Use by Children and Youth with Mental Health Conditions: A Health Equity Agenda. Community Ment Health J [Internet]. 2022 Oct 17;58(7):1225–39. Available from: https://link.springer.com/10.1007/s10597-022-00937-7

17. Fullen BM, Baxter GD, O’Donovan BGG, Doody C, Daly L, Hurley DA. Doctors’ attitudes and beliefs regarding acute low back pain management: A systematic review. Pain [Internet]. 2008;136(3):388–96. Available from: https://www.sciencedirect.com/science/article/pii/S0304395908000122

18. Claudius I, Donofrio JJ, Lam CN, Santillanes G. Impact of Boarding Pediatric Psychiatric Patients on a Medical Ward. Hosp Pediatr [Internet]. 2014 May 1;4(3):125–32. Available from: 10.1542/hpeds.2013-0079

19. Case BG, Olfson M, Marcus SC, Siegel C. Trends in the Inpatient Mental Health Treatment of Children and Adolescents in US Community Hospitals Between 1990 and 2000. Arch Gen Psychiatry [Internet]. 2007 Jan 1;64(1):89–96. Available from: 10.1001/archpsyc.64.1.89

20. Levine LJ, Schwarz DF, Argon J, Mandell DS, Feudtner C. Discharge Disposition of Adolescents Admitted to Medical Hospitals After Attempting Suicide. Arch Pediatr Adolesc Med [Internet]. 2005 Sep 1;159(9):860–6. Available from: 10.1001/archpedi.159.9.860

21. Smith WG, Collings A, Degraaf A. Young people admitted on a Form 1 to a general hospital: A worrisome trend. Paediatr Child Health [Internet]. 2004 Apr 1;9(4):228–34. Available from: 10.1093/pch/9.4.228

22. Mansbach JM, Wharff E, Austin SB, Ginnis K, Woods ER. Which Psychiatric Patients Board on the Medical Service? Pediatrics [Internet]. 2003 Jun 1;111(6):e693–8. Available from: https://publications.aap.org/pediatrics/article/111/6/e693/28595/Which-Psychiatric-Patients-Board-on-the-Medical

23. Valdivia M, Ebner D, Fierro V, Gajardo C, Miranda R. Hospitalización por intento de suicidio en población pediátrica: una revisión de cuatro años. Rev Chil Neuropsiquiatr [Internet]. 2001 Sep;39(3). Available from: http://www.scielo.cl/scielo.php?script=sci_arttext&pid=S0717-92272001000300005&lng=en&nrm=iso&tlng=en

24. Gasquet I, Choquet M. Hospitalization in a pediatric ward of adolescent suicide attempters admitted to general hospitals. J Adolesc Heal [Internet]. 1994;15(5):416–22. Available from: https://www.sciencedirect.com/science/article/pii/1054139X94902674

25. Kölch M, Reis O, Ulbrich L, Schepker R. Mental disorders in minors during the COVID-19 pandemic. Dtsch Arztebl Int [Internet]. 2023 May 19; Available from: https://www.aerzteblatt.de/10.3238/arztebl.m2023.0010

26. Duarte V, Zelaya L. Caracterización clínica y demográfica de la hospitalización psiquiátrica infantojuvenil en un hospital general. Pediatría (Asunción) [Internet]. 2019 Jul 30;46(2):90–6. Available from: https://revistaspp.org/index.php/pediatria/article/view/496

27. Plemmons G, Hall M, Doupnik S, Gay J, Brown C, Browning W, et al. Hospitalization for Suicide Ideation or Attempt: 2008–2015. Pediatrics [Internet]. 2018 Jun 1;141(6):e20172426. Available from: 10.1542/peds.2017-2426

28. Wallis M, Akhtar F, Azam M. Emergency Admissions of Children and Young People with Mental Health Needs to the Paediatric Ward [Internet]. Vol. 111, Irish medical journal. Department of Paediatrics, Wexford General Hospital, Ireland; 2018. p. 795. Available from: http://europepmc.org/abstract/MED/30520288

29. Santillanes G, Kearl Y, Lam C, Claudius I. Involuntary Psychiatric Holds in Preadolescent Children. West J Emerg Med [Internet]. 2017 Oct 18;18(6):1159–65. Available from: http://www.escholarship.org/uc/item/8vc737vg

30. Suetani S, Yiu SM, Batterham M. Defragmenting paediatric anorexia nervosa: the Flinders Medical Centre Paediatric Eating Disorder Program. Australas Psychiatry [Internet]. 2015;23(3):245–8. Available from: 10.1177/1039856215579543

31. Chang Y-S, Liao F-T, Huang L-C, Chen S-L. The Treatment Experience of Anorexia Nervosa in Adolescents from Healthcare Professionals’ Perspective: A Qualitative Study. Int J Environ Res Public Health [Internet]. 2023 Jan 1;20(1):794. Available from: https://www.mdpi.com/1660-4601/20/1/794

32. Ramritu P, Courtney M, Stanley T, Finlayson K. Experiences of the Generalist Nurse Caring for Adolescents with Mental Health Problems. J Child Heal Care [Internet]. 2002 Dec 1;6(4):229– 44. Available from: 10.1177/136749350200600401

33. King SJ, Turner de S. Caring for adolescent females with anorexia nervosa: registered nurses’ perspective. J Adv Nurs [Internet]. 2000 Jul;32(1):139–47. Available from: http://doi.wiley.com/10.1046/j.1365-2648.2000.01451.x

34. Lakeman R, McIntosh C. Perceived confidence, competence and training in evidence-based treatments for eating disorders: a survey of clinicians in an Australian regional health service. Australas Psychiatry [Internet]. 2018;26(4):432–6. Available from: 10.1177/1039856218766124

35. Wu W, Chen S. Nurses’ perceptions on and experiences in conflict situations when caring for adolescents with anorexia nervosa: A qualitative study. Int J Ment Health Nurs [Internet]. 2021 Oct 27;30(S1):1386–94. Available from: https://onlinelibrary.wiley.com/doi/10.1111/inm.12886

36. Hampton E, Richardson JE, Bostwick S, Ward MJ, Green C. The Current and Ideal State of Mental Health Training: Pediatric Resident Perspectives. Teach Learn Med [Internet]. 2015 Apr 3;27(2):147–54. Available from: http://www.tandfonline.com/doi/full/10.1080/10401334.2015.1011653

37. Ramjan LM, Gill BI. Original Research: An Inpatient Program for Adolescents with Anorexia Experienced as a Metaphoric Prison. AJN Am J Nurs [Internet]. 2012;112(8). Available from: https://journals.lww.com/ajnonline/Fulltext/2012/08000/Original_Research__An_Inpatient_Program_for.20.aspx

38. Buckley S. Caring for those with mental health conditions on a children’s ward. Br J Nurs [Internet]. 2010 Oct 1;19(19):1226–30. Available from: http://www.magonlinelibrary.com/doi/10.12968/bjon.2010.19.19.79303

39. Happell B, Moxham L, Reid-Searl K, Dwyer T, Kahl J, Morris J, et al. Promoting mental health care in a rural paediatric unit through participatory action research. Aust J Rural Health [Internet]. 2009 Jun;17(3):155–60. Available from: https://onlinelibrary.wiley.com/doi/10.1111/j.1440-1584.2009.01061.x

40. Watson E. CAMHS liaison: supporting care in general paediatric settings. Paediatr Nurs [Internet]. 2006 Feb;18(1):30–3. Available from: https://www.proquest.com/scholarly-journals/camhs-liaison-supporting-care-general-paediatric/docview/218919947/se-2?accountid=14511

41. Anderson M, Standen P, Noon J. Nurses’ and doctors’ perceptions of young people who engage in suicidal behaviour: a contemporary grounded theory analysis. Int J Nurs Stud [Internet]. 2003 Aug;40(6):587–97. Available from: https://linkinghub.elsevier.com/retrieve/pii/S0020748903000543

42. Worsley D, Barrios E, Shuter M, Pettit AR, Doupnik SK. Adolescents’ Experiences During “Boarding” Hospitalization While Awaiting Inpatient Psychiatric Treatment Following Suicidal Ideation or Suicide Attempt. Hosp Pediatr [Internet]. 2019 Nov 1;9(11):827–33. Available from: https://publications.aap.org/hospitalpediatrics/article/9/11/827/26717/Adolescents-Experiences-During-Boarding

43. Todd E, Adamson R, Leith E, Davies A. Mind the gap: addressing mental health competency on the acute paediatric ward. Arch Dis Child - Educ Pract Ed [Internet]. 2023 Feb 9;edpract-2022-324106. Available from: https://ep.bmj.com/lookup/doi/10.1136/archdischild-2022-324106

44. Bolland R, Richardson J, Calnan R. How professionals should communicate with children who have mental healthcare needs. Nurs Child Young People. 2017;29(1):20–4.

