## Supplementary file_Quality assessment for "‘Admissions to paediatric medical wards with a primary mental health diagnosis: a systematic review of the literature’"

Table S1. CASP Qualitative Studies Checklist*

|  |  | Criteria for qualitative studies | | | | | | | | | |  |
| --- | --- | --- | --- | --- | --- | --- | --- | --- | --- | --- | --- | --- |
| # | Authors | 1 | 2 | 3 | 4 | 5 | 6 | 7 | 8 | 9 | 10 | Assessment |
| 1 | Chang et al (2023) | Y | Y | Y | Y | Y | C | Y | Y | Y | Y | High |
| 2 | Wu and Chen (2021) | Y | Y | Y | Y | Y | C | Y | Y | Y | Y | High |
| 3 | Worsley (2019) | Y | Y | N | Y | Y | N | C | Y | Y | Y | High |
| 4 | Hampton et al (2015) | Y | Y | Y | Y | Y | Y | Y | Y | Y | Y | High |
| 5 | Ramjan and Gil (2012) | Y | Y | C | Y | Y | N | Y | C | Y | C | Moderate |
| 6 | Happell et al (2009) | Y | Y | Y | Y | Y | C | Y | N | Y | Y | High |
| 7 | Watson (2006) | Y | Y | N | N | Y | N | N | C | Y | C | Low |
| 8 | Anderson et al (2003) | Y | Y | Y | C | Y | N | C | Y | Y | Y | High |
| 9 | King and Turner (2000) | Y | Y | Y | C | Y | N | Y | C | Y | C | Moderate |

*CASP criteria for qualitative studies: 1. Was there a clear statement of the aims of the research?; 2. Was a qualitative methodology appropriate?; 3. Was the research design appropriate to address the aims of the research?; 4. Was the recruitment strategy appropriate to the aims of the research?; 5. Was the data collected in a way that addressed the research issue?; 6. Has the relationship between researcher and participants been adequately considered?; 7 Have ethical issues been considered?; 8. Was the data analysis sufficiently rigorous?; 9. Is there a clear statement of the findings?; 10. How valuable is the research? (Y: Yes, N: No, C: Can’t tell)

**Table S2. AXIS scale for cross-sectional studies***

|  | | Kölch et al (2023) | Ibeziako et al (2022) | Duarte and Zelaya (2019) | Plemmons et al (2018) | Wallis (2018) | Gallagher et al (2017) | Santillanes et al (2017) | Claudius et al (2014) | Case et al (2007) | Levine et al (2005) | Smith et al (2014) | Mansbach et al (2003) | Ramritu et al (2002) | Valdivia et al (2001­) | Gasquet and Choquet (1994) |
| --- | --- | --- | --- | --- | --- | --- | --- | --- | --- | --- | --- | --- | --- | --- | --- | --- |
| Introduction | Were the aims/objectives of the study clear? | Y | Y | Y | Y | Y | Y | Y | Y | Y | Y | Y | Y | Y | Y | Y |
| Methods | Was the study design appropriate for the stated aim(s)? | Y | Y | Y | Y | Y | Y | Y | Y | Y | Y | Y | Y | Y | Y | Y |
|  | Was the sample size justified? | N | N | Y | Y | N | N | N | N | Y | Y | N | N | N | N | N |
|  | Was the target/reference population clearly defined? (Is it clear who the research was about?) | N | Y | Y | Y | Y | Y | Y | Y | Y | Y | Y | Y | Y | Y | Y |
|  | Was the sample frame taken from an appropriate population base so that it closely represented the target/reference population under investigation? | Y | Y | Y | Y | Y | Y | Y | Y | Y | Y | Y | Y | Y | Y | Y |
|  | Was the selection process likely to select subjects/participants that were representative of the target/reference population under investigation? | Y | Y | Y | Y | Y | Y | Y | Y | Y | Y | Y | Y | Y | Y | Y |
|  | Were measures undertaken to address and categorize non-responders? | N | N | N | N | N | N | N | N | N | N | N | Y | N | N | N |
|  | Were the risk factor and outcome variables measured appropriate to the aims of the  study? | Y | Y | Y | Y | Y | Y | Y | Y | Y | Y | Y | Y | Y | Y | Y |
|  | Were the risk factor and outcome variables measured correctly using instruments/measurements that had been trialled, piloted or published previously? | Y | Y | Y | Y | Y | Y | Y | Y | Y | Y | Y | Y | Y | Y | Y |
|  | Is it clear what was used to determined statistical significance and/or precision estimates? (e.g., p values, CIs) | Y | Y | NA | Y | N | N | Y | N | Y | Y | N | Y | NA | NA | N |
|  | Were the methods (including statistical methods) sufficiently described to enable them to be repeated? | Y | Y | Y | Y | N | Y | Y | Y | Y | Y | N | N | N | N | Y |
| Results | Were the basic data adequately described? | Y | Y | Y | Y | Y | Y | Y | Y | Y | Y | Y | Y | Y | Y | Y |
|  | Does the response rate raise concerns about non-response bias? | ND | ND | ND | ND | ND | ND | ND | ND | ND | ND | ND | N | ND | N | N |
|  | If appropriate, was information about no responders described? | N | N | N | N | N | N | N | N | N | N | N | Y | N | N | N |
|  | Were the results internally consistent? | Y | Y | Y | Y | Y | Y | Y | Y | Y | Y | Y | Y | Y | Y | Y |
|  | Were the results for the analyses described in the methods, presented? | N | Y | Y | Y | N | Y | Y | Y | Y | Y | Y | Y | Y | Y | Y |
| Discussion | Were the authors’ discussions and conclusions justified by the results? | Y | Y | Y | Y | N | Y | Y | Y | Y | Y | Y | Y | Y | Y | Y |
|  | Were the limitations of the study discussed? | Y | Y | Y | Y | N | Y | Y | Y | Y | Y | N | Y | N | N | N |
| Others | Were there any funding sources or conflicts of interest that may affect the authors’ interpretation of the results? | NDis | N | NDis | N | NDis | N | N | N | NDis | NDis | NDis | NDis | NDis | NDis | NDis |
|  | Was ethical approval or consent of participants attained? | NS | Y | Y | Y | NS | Y | Y | Y | NS | Y | Y | Y | Y | NS | NS |

*The tool does not provide a numerical scale for assessing the quality of the study, it has areas to record assessment using “Yes”, “No” or “Don’t Know/comments” answer for each of the 20 questions.

Abbreviations: Y: Yes; N: No; DK: Don’t know; Comments: not described (ND), not disclosed (NDis), not stated (NS), not applicable (NA)
