## Supplementary material for "‘Admissions to paediatric medical wards with a primary mental health diagnosis: a systematic review of the literature’": Supplmentary file_PROSPERO

### Systematic review

A list of fields that can be edited in an update can be found here

#### 1. * Review title.

Give the title of the review in English

A rapid review of current literature on admissions to paediatric wards with a primary mental health diagnosis

#### 2. Original language title.

For reviews in languages other than English, give the title in the original language. This will be displayed with the English language title.

#### 3. * Anticipated or actual start date.

Give the date the systematic review started or is expected to start.

08/08/2022

#### 4. [2 changes] * Anticipated completion date.

Give the date by which the review is expected to be completed.

30/04/2023

#### 5. [3 changes] * Stage of review at time of this submission.

**This field uses answers to initial screening questions. It cannot be edited until after registration.**

Tick the boxes to show which review tasks have been started and which have been completed.

Update this field each time any amendments are made to a published record.

The review has not yet started: No

| Review stage | Started | Completed |
| --- | --- | --- |
| Preliminary searches | Yes | Yes |
| Piloting of the study selection process | Yes | Yes |
| Formal screening of search results against eligibility criteria | Yes | Yes |
| Data extraction | Yes | Yes |
| Risk of bias (quality) assessment | Yes | Yes |
| Data analysis | Yes | Yes |

Provide any other relevant information about the stage of the review here.

The completed manuscript was submitted to 'Archives of Disease in Childhood (BMJ)' on the 8th of November 2023. The title of the manuscript is: ‘Admissions to paediatric medical wards with a primary mental health diagnosis: a systematic review of the literature’.

Moreover, for quantitative studies, the Appraisal tool for Cross-Sectional Studies (AXIS) was used to assess quality.

The completed manuscript was submitted to 'Archives of Disease in Childhood (BMJ)' on the 8th of November 2023. The title of the manuscript is: ‘Admissions to paediatric medical wards with a primary mental health diagnosis: a systematic review of the literature’.

Moreover, for quantitative studies, the Appraisal tool for Cross-Sectional Studies (AXIS) was used to assess quality.

#### 6. * Named contact.

The named contact is the guarantor for the accuracy of the information in the register record. This may be any member of the review team.

Adriana del Pilar Vazquez Vazquez

#### Email salutation (e.g. "Dr Smith" or "Joanne") for correspondence:

Dr Vazquez Vazquez

#### 7. * Named contact email.

Give the electronic email address of the named contact.

#### 8. Named contact address

Give the full institutional/organisational postal address for the named contact.

UCL Great Ormond Street Institute of Child Health, 30 Guilford St, London WC1N 1EH

#### 9. Named contact phone number.

Give the telephone number for the named contact, including international dialling code.

#### 10. * Organisational affiliation of the review.

Full title of the organisational affiliations for this review and website address if available. This field may be completed as 'None' if the review is not affiliated to any organisation.

Population, Policy and Practice Research Programme, UCL Great Ormond Street Institute of Child Health, London, UK

#### Organisation web address:

https://www.ucl.ac.uk/child-health/great-ormond-street-institute-child-health-0

#### [1 change] 11. * Review team members and their organisational affiliations.

Give the personal details and the organisational affiliations of each member of the review team. Affiliation refers to groups or organisations to which review team members belong. **NOTE: email and country now MUST be entered for each person, unless you are amending a published record.**

Dr Adriana del Pilar Vazquez Vazquez. Population, Policy and Practice Research Programme, UCL Great Ormond Street Institute of Child Health, London, UK
Miss Abigail Smith. Population, Policy and Practice Research Programme, UCL Great Ormond Street Institute of Child Health, London, UK
Professor Faith Gibson. University of Surrey, Guildford, UK
Professor Helen Roberts. Population, Policy and Practice Research Programme, UCL Great Ormond Street Institute of Child Health, London, UK
Dr Gabrielle Mathews. CYP Transformation Team, NHS England and NHS Improvement London, London, UK
Dr Joseph Ward. Population, Policy and Practice Research Programme, UCL Great Ormond Street Institute of Child Health, London, UK
Professor Russell Viner. Population, Policy and Practice Research Programme, UCL Great Ormond Street Institute of Child Health, London, UK
Dr Dasha Nicholls. Imperial College London, Faculty of Medicine, Department of Brain Sciences, London, UK
Dr Francesca Cornaglia. Queen Mary University of London, London, UK
Dr Damian Roland. University Hospitals of Leicester NHS Trust, Leicester, UK
Miss Kirsty Phillips. Population, Policy and Practice Research Programme, UCL Great Ormond Street Institute of Child Health, London, UK
Dr Lee Hudson. Population, Policy and Practice Research Programme, UCL Great Ormond Street Institute of Child Health, London, UK

#### 12. * Funding sources/sponsors.

Details of the individuals, organizations, groups, companies or other legal entities who have funded or sponsored the review.

The review is being undertaken as part of a wider project (MAPS: Mental Health Admissions to Paediatric Wards Study) that has been funded by the National Institute for Health and Care Research.

#### Grant number(s)

State the funder, grant or award number and the date of award

NIHR135036

#### 13. * Conflicts of interest.

List actual or perceived conflicts of interest (financial or academic).

None

#### 14. Collaborators. [1 change]

Give the name and affiliation of any individuals or organisations who are working on the review but who are not listed as review team members. **NOTE: email and country must be completed for each person, unless you are amending a published record.**

#### 15. * Review question.

State the review question(s) clearly and precisely. It may be appropriate to break very broad questions down into a series of related more specific questions. Questions may be framed or refined using PI(E)COS or similar where relevant.

1. What is the trend in numbers of admissions/temporal changes in numbers of admissions of children admitted to paediatric wards or adult wards because of a primary mental health diagnosis?

2. What are the risk factors for adverse care/adverse outcomes for children, young people and families during admissions to paediatric wards (or adult general wards) because of a primary mental health diagnosis?

3. What are the reported experiences of children, young people and their families during admissions to paediatric wards (or adult general wards) because of a primary mental health diagnosis?

4. What are the reported experiences of clinical staff on paediatric wards (or adult general wards) during the admissions of children and young people admitted because of a primary mental health diagnosis?

5. Is there evidence of interventions or quality improvement projects aimed at improving the care of children, young people and families during admissions to paediatric wards (or adult wards) because of a primary mental health diagnosis?

#### 16. * Searches.

State the sources that will be searched (e.g. Medline). Give the search dates, and any restrictions (e.g. language or publication date). Do NOT enter the full search strategy (it may be provided as a link or attachment below.)

We will search across four databases (PubMed, Embase, PsycINFO and Web of Science) and an additional search of Google Scholar to identify unpublished data or additional studies from 1990 onwards. We will include studies published in all languages.

#### 17. URL to search strategy.

Upload a file with your search strategy, or an example of a search strategy for a specific database, (including the keywords) in pdf or word format. In doing so you are consenting to the file being made publicly accessible. Or provide a URL or link to the strategy. Do NOT provide links to your search **results**.

https://www.crd.york.ac.uk/PROSPEROFILES/350655_STRATEGY_20220801.pdf

Alternatively, upload your search strategy to CRD in pdf format. Please note that by doing so you are consenting to the file being made publicly accessible.

Do not make this file publicly available until the review is complete

#### 18. * Condition or domain being studied.

Give a short description of the disease, condition or healthcare domain being studied in your systematic review.

This rapid review is essential to synthesise the evidence available about the trends in admissions for primary mental health reasons to acute paediatric wards or adult general wards, the reasons for admissions, the contributing factors to the admissions, the clinical needs of children and young people when admitted, and the views/experiences of children and young people, families and health professionals to understand the context of care.

#### 19. * Participants/population.

Specify the participants or populations being studied in the review. The preferred format includes details of both inclusion and exclusion criteria.

Inclusion: studies with participants less than or equal to 18 years old admitted to any paediatric ward or adult general wards. Exclusion: studies with participants who are over 18 years old and/or admitted to specialist mental health wards.

#### 20. * Intervention(s), exposure(s).

Give full and clear descriptions or definitions of the interventions or the exposures to be reviewed. The preferred format includes details of both inclusion and exclusion criteria.

Admission to an acute paediatric ward or adult general wards because of a primary mental health diagnosis.

#### 21. * Comparator(s)/control. [1 change]

Where relevant, give details of the alternatives against which the intervention/exposure will be compared (e.g. another intervention or a non-exposed control group). The preferred format includes details of both inclusion and exclusion criteria.

Our team will compare different types/reasons for admission to an acute paediatric ward or adult general wards because of a primary mental health diagnosis.

#### 22. * Types of study to be included.

Give details of the study designs (e.g. RCT) that are eligible for inclusion in the review. The preferred format includes both inclusion and exclusion criteria. If there are no restrictions on the types of study, this should be stated.

Inclusions: Systematic reviews on any of the five review questions; Quantitative studies: Observational studies in the case of the epidemiology of numbers of admissions, quality improvement studies, randomized and non-randomized controlled trials; Qualitative studies addressing review questions 2-5; Reports by professional bodies published on any of the above five review questions containing unique data.

Exclusion: Non-systematic reviews (however a snowballing approach will be taken using reference lists); Studies which only study emergency department attendances (thus in included studies children and young people must have had to be admitted to a paediatric ward); Studies where it is not possible to disaggregate data for children and young people from adults.

#### 23. Context.

Give summary details of the setting or other relevant characteristics, which help define the inclusion or exclusion criteria.

No restrictions based on the geographical location of the articles considered

#### 24. * Main outcome(s).

Give the pre-specified main (most important) outcomes of the review, including details of how the outcome is defined and measured and when these measurement are made, if these are part of the review inclusion criteria.

1. Establishing the trends in admissions and characteristics of the admissions (sociodemographic factors, diagnoses and reasons admitted).

2. Establishing the factors that influence decisions to admit children and young people to paediatric wards for primary mental health problems.

3. Establishing the views/experiences of children and young people, families and health care professionals during admissions.

#### Measures of effect

Please specify the effect measure(s) for you main outcome(s) e.g. relative risks, odds ratios, risk difference, and/or 'number needed to treat.

#### 25. * Additional outcome(s).

List the pre-specified additional outcomes of the review, with a similar level of detail to that required for main outcomes. Where there are no additional outcomes please state 'None' or 'Not applicable' as appropriate to the review

None

#### Measures of effect

Please specify the effect measure(s) for you additional outcome(s) e.g. relative risks, odds ratios, risk difference, and/or 'number needed to treat.

#### 26. * Data extraction (selection and coding).

Describe how studies will be selected for inclusion. State what data will be extracted or obtained. State how this will be done and recorded.

Titles and abstracts of all papers retrieved will be screened by two reviewers, who independently will identify studies that met all inclusion criteria and none of the exclusion criteria. Disagreements that cannot be resolved via consensus will be reviewed independently by another author who had not participated in the screening. All full-text studies meeting initial criteria will be reviewed by two reviewers for final inclusion in the rapid review.

Two reviewers will extract data from included studies using a data collection form. We will collect data on the following variables: author, country of study, start and end dates, type of study, study design, healthcare setting, sample size, population and results (related to the five review questions).

The results of the search will be imported into Covidence, an online software tool for systematic reviews. Duplicate records will be removed using Covidence.

National Institute for Health Research

#### 27. * Risk of bias (quality) assessment.

State which characteristics of the studies will be assessed and/or any formal risk of bias/quality assessment tools that will be used.

The final step in the data handling process is quality control of the included studies using a combination of different quality assessment tools. For qualitative studies, the CASP (Critical Appraisal Skills Programme) checklists will be used and for observational studies, the Newcastle-Ottawa scale will be used to assess the risk of bias. For interventions: for non-randomized studies the ROBINS-1 will be used, for randomized, controlled studies Cochrane's risk tool (RoB) will be used.

#### 28. * Strategy for data synthesis. [1 change]

Describe the methods you plan to use to synthesise data. This **must not be generic text** but should be **specific to your review** and describe how the proposed approach will be applied to your data. If meta-analysis is planned, describe the models to be used, methods to explore statistical heterogeneity, and software package to be used.

According to the experience of the research team, it is anticipated that there will not be sufficient studies for quantitative synthesis.

Therefore, data will be summarized in tables in a tabular and narrative format.

We propose the following structure:

1. Summary of included systematic reviews, and of any quantitative studies that cover the epidemiology of numbers of admissions, and randomized and non-randomized controlled trials.

2. Summary of qualitative interventions that aimed to improve the care of children, young people and families during admissions to paediatric wards (or adult wards) because of a primary mental health diagnosis.

3. Summary of the common views, experiences, feelings, and perceptions of children, young people and their families during admissions to paediatric wards (or adult general wards) because of a primary mental health diagnosis.

4. Summary of the common views, experiences, feelings, and perceptions of clinical staff on paediatric wards (or adult general wards) during the admissions of children and young people admitted because of a primary mental health diagnosis.

5. Summary of any tools commonly used for deciding admission to an acute paediatric ward or adult general ward because of a primary mental health diagnosis.

6. Summary of the quality of the systematic reviews and possible sources of bias.

7. Summary of any knowledge gap which has been identified.

#### 29. * Analysis of subgroups or subsets.

State any planned investigation of 'subgroups'. Be clear and specific about which type of study or participant will be included in each group or covariate investigated. State the planned analytic approach.

None planned.

#### 30. * Type and method of review.

Select the type of review, review method and health area from the lists below.

##### Type of review

Cost effectiveness

No

Diagnostic

No

Epidemiologic

No

Individual patient data (IPD) meta-analysis

No

Intervention

No

Living systematic review

No

Meta-analysis

No

Methodology

No

Narrative synthesis

Yes

Network meta-analysis
No

Pre-clinical
No

Prevention
No

Prognostic
No

Prospective meta-analysis (PMA)
No

Review of reviews
No

Service delivery
No

Synthesis of qualitative studies
No

Systematic review
Yes

Other
No

#### Health area of the review

Alcohol/substance misuse/abuse
No

Blood and immune system
No

Cancer
No

Cardiovascular
No

Care of the elderly
No

Child health
Yes

Complementary therapies

No

COVID-19

No

Crime and justice

No

Dental

No

Digestive system

No

Ear, nose and throat

No

Education

No

Endocrine and metabolic disorders

No

Eye disorders

No

General interest

No

Genetics

No

Health inequalities/health equity

No

Infections and infestations

No

International development

No

Mental health and behavioural conditions

Yes

Musculoskeletal

No

Neurological

No

Nursing

No

Obstetrics and gynaecology

No

Oral health

No

Palliative care

No

Perioperative care

No

Physiotherapy

No

Pregnancy and childbirth

No

Public health (including social determinants of health)

Yes

Rehabilitation

No

Respiratory disorders

No

Service delivery

No

Skin disorders

No

Social care

No

Surgery

No

Tropical Medicine

No

Urological

No

Wounds, injuries and accidents

No

Violence and abuse

No

#### 31. Language.

Select each language individually to add it to the list below, use the bin icon  to remove any added in error.

English

There is an English language summary.

#### 32. * Country.

Select the country in which the review is being carried out. For multi-national collaborations select all the countries involved.

England

#### 33. Other registration details.

Name any other organisation where the systematic review title or protocol is registered (e.g. Campbell, or The Joanna Briggs Institute) together with any unique identification number assigned by them. If extracted data will be stored and made available through a repository such as the Systematic Review Data Repository (SRDR), details and a link should be included here. If none, leave blank.

#### [1. change] 34. Reference and/or URL for published protocol.

If the protocol for this review is published provide details (authors, title and journal details, preferably in Vancouver format)

Add web link to the published protocol.

Or, upload your published protocol here in pdf format. Note that the upload will be publicly accessible.

No I do not make this file publicly available until the review is complete

Please note that the information required in the PROSPERO registration form must be completed in full even if access to a protocol is given.

#### 35. Dissemination plans.

Do you intend to publish the review on completion?

Yes

Give brief details of plans for communicating review findings.?

#### 36. Keywords.

Give words or phrases that best describe the review. Separate keywords with a semicolon or new line. Keywords help PROSPERO users find your review (keywords do not appear in the public record but are included in searches). Be as specific and precise as possible. Avoid acronyms and abbreviations unless these are in wide use.

#### 37. Details of any existing review of the same topic by the same authors.

If you are registering an update of an existing review give details of the earlier versions and include a full bibliographic reference, if available.

#### 38. * Current review status. [1 change]

Update review status when the review is completed and when it is published.New registrations must be ongoing so this field is not editable for initial submission.

Please provide anticipated publication date

Review_Completed_not_published

#### 39. Any additional information.

Provide any other information relevant to the registration of this review.

#### 40. Details of final report/publication(s) or preprints if available. [1 change]

Leave empty until publication details are available OR you have a link to a preprint (NOTE: this field is not editable for initial submission). List authors, title and journal details preferably in Vancouver format.

Give the link to the published review or preprint.
